# Incidence of Thirty-day MACE among patients presenting to emergency department with low-risk chest pain in a tertiary care hospital: A prospective study

**DOI:** 10.1101/2022.05.17.22275224

**Authors:** Sanwar Khokhar, Abhishek Jaiswal, Raman Abhi, Mohammed Hasnain Reza

## Abstract

**Background:** Current guidelines for low-risk chest pain patients recommend obtaining serial ECGs and serial measurements of cardiac troponin between 6 and 12 hours. As a result, the majority of patients require prolonged assessment before safe discharge. There is a need to identify these patients promptly to help in reducing the time to provide the treatment as well as reduce the burden over the ED. Present study was done with the objective of estimating the incidence of thirty-day Major Averse Cardiac Event (MACE) in patients presenting to emergency department with low-risk chest pain, and to compare the Thrombolysis In Myocardial Infarction (TIMI), HEART and Emergency Department Assessment of Chest Pain Score (EDACS) Score in patients with low-risk chest pain.

**Methods:** Present study was descriptive follow up study done at a tertiary care hospital (Fortis Memorial Research Institute, in Gurugram, Haryana, India. Study was conducted from Jan 2018 to Jan 2019. All the patient reporting with low-risk chest pain during study period were recruited in the study. Semi-structured interview schedule was used for the data collection. Outcome variable was MACE (Major adverse cardiac event) event in 30 days.

**Results:** Total 156 participants were included in the study. Mean age of participants was 44.1 years. Out of 156 participants, 10 (6.4%) reported MACE in 30 days of presentation. We found that HEART and EDACS score had incidence of MACE less than 2% in their low-risk groups and TIMI score had incidence of MACE >2% in its low-risk group.

**Conclusion:** EDACS and HEART score can be used in the Emergency department to identify the low-risk chest pain patients. This could help in early identification and save time and other resources.

**What is already known on this topic:** Current guidelines for low-risk chest pain patients recommend obtaining serial ECGs and serial measurements of (non-high sensitivity) cardiac troponin between 6 and 12 hours after patient presentation to the ED. As a result, the majority of patients require prolonged assessment before safe discharge. Prolonged assessment leads to increased health care costs and ED crowding, which has been shown to lead to increased adverse events in patients with both acute and non-acute coronary syndrome–related chest pain. The efficient identification of low-risk patients who can be safely discharged after rapid assessment in the ED remains an important issue. Risk assessment scores have been developed for chest pain, among these few are TIMI score, Heart score, and EDACS score.

**What this study adds:** Overall incidence of 30-day MACE was less 10% among the patients presenting to emergency department of FMRI Gurugram, Haryana with low-risk chest pain. HEART, and EDACS scores performed better in identifying the low-risk category than the TIMI score. Among these EDACS was the best, with none of the participants in low-risk category having 30-day MACE.

**How this study might affect research, practice or policy:** EDACS and HEART score can be used in the Emergency department to identify the low-risk chest pain patients. This could help in early identification and save time and other resources.

## Introduction

Chest pain or Chest discomfort or Chest uneasiness is second most common reasons for emergency department (ED) visit, accounting for approximately 4.9% of the total ED visits yearly. (1) Given that Asian Indians have a mean onset of coronary artery disease (CAD) 5–10 years earlier than the western world, the burden of chest pain visits to EDs in India is likely much higher. (2)

Current guidelines for low-risk chest pain patients (3) recommend obtaining serial ECGs and serial measurements of (non-high sensitivity) cardiac troponin between 6 and 12 hours after patient presentation to the ED. As a result, the majority of patients require prolonged assessment before safe discharge despite that only 15% to 25% of them receive a final diagnosis of acute coronary syndrome. (4) This prolonged assessment leads to increased health care costs (5) and ED crowding, which has been shown to lead to increased adverse events in patients with both acute and non-acute coronary syndrome–related chest pain. (6) The efficient identification of low-risk patients who can be safely discharged after rapid assessment in the ED remains an important issue. And early discharge also has a risk up to 2-5% of patients with ACS are inappropriately discharged from emergency department every year. (7)

Risk assessment scores have been developed for chest pain, among these few are TIMI score (8), Heart score (9), and EDACS score (10).

Present study was done with the objective of estimating the incidence of thirty-day Major Averse Cardiac Event (MACE) in patients presenting to emergency department with low-risk chest pain, and to compare the Thrombolysis In Myocardial Infarction (TIMI), HEART and Emergency Department Assessment of Chest Pain Score (EDACS) Score in patients with low-risk chest pain.

## Methodology

Present study was descriptive follow up study done at a tertiary care hospital (Fortis Memorial Research Institute (FMRI)), in Gurugram, Haryana, India. Patients presenting with chest pain or chest discomfort or chest uneasiness or chest heaviness reporting to department of emergency and trauma department of FMRI hospital were recruited in the study if they fulfilled the inclusion and exclusion criteria and gave consent for the study. Inclusion criteria: a) Patient presents with chest pain, b) patient age >18 years, c) patient should be able to communicate. Exclusion criteria: a) Refusal to give consent, b) positive troponin value, and c) ST segment changes in ECG.

Study was conducted from Jan 2018 to Jan 2019. Each participant was followed for thirty days of emergency visit via phone or patient’s hospital visit. All the patient reporting with low-risk chest pain during study period were recruited in the study. Semi-structured interview schedule was used for the data collection having questions on sociodemographic variable, chest pain, smoking tobacco, obesity, family history of cardiac illness, previous diagnosis of hypertension or diabetic or coronary artery diseases, ECG findings, Troponin I level. Outcome variable was MACE event in 30 days.

### Operational definitions

a. Low Risk Chest Pain (3): Low risk chest pain is defined as patient complaining of chest pain or chest discomfort or chest uneasiness or chest heaviness with no hemodynamic derangements or arrhythmias, a normal or near normal electrocardiogram (ECG), negative initial cardiac injury markers.
b. Thirty-day MACE (10,11) : It is defined as development of non-ST-elevation myocardial infarction (NSTEMI) or STEMI or emergency revascularization or cardiovascular death or cardiac arrest or cardiogenic shock or high-grade atrioventricular block within a 30-day period

For STEMI AHA standard definition was used. (12)

TIMI Score Calculation (8) - According to the TIMI score patients are divided into low (score 0-1), intermediate (score 2-4) and high (score 5-7) risk categories. Each of the following criteria constitutes one point for TIMI scoring: a) Age ≥65 years, b) Three or more risk factors for coronary artery disease (CAD) (family history of CAD, hypertension, hypercholesterolemia, diabetes mellitus, tobacco use), c) Known CAD (stenosis >50%), d) Aspirin use in the past 7 days, e) Severe angina (≥2 episodes in 24 hours), f) ST deviation ≥0.5 mm, g) elevated cardiac marker level

Heart Score Calculation (9) – Scores are given as 0, 1, or 2 on following points, a) history, b) ECG, c) Age, d) Risk factors, e) Troponin. The HEART score divides patients into low (0-3), intermediate (4-6) or high-risk groups (7-10), with mean risks of an event of 0.9%, 12% and 65%, respectively.

EDACS Score (10) – In EDACS Score low score is identified as score <16.

As few of the scoring systems have questions based on history of participants recall bias can be there. To circumvent this, all these questions were probed and relevant information to confirm these histories based on reports and prescription was done.

### Sample size

Complete enumeration. All the participants reporting to the ED with acute chest pain and fulfilling the exclusion and inclusion criteria were recruited in the study after taking written informed consent.

Statistical analysis: Data entry was done in Microsoft excel. Categorical variables were reported as frequency and percentages. Continuous variables were reported as mean and standard deviation. Incidence of 30-day MACE was reported as proportion with 95% C.I. Statistical analysis was done using STATA 16 (StataCorp. 2019. Stata Statistical Software: Release 16).

### Ethics

Written informed consent was taken from all the participants after informing about the study objective. Ethical permission for the study was taken from Hospitals Ethics Committee (FMRI, Gurugram) (IEC code no.: 2018-007TH-22).

## Patient and Public Involvement

It was not appropriate or possible to involve patients or the public in the design, or conduct, or reporting, or dissemination plans of our research. Public were not involved in the design, or conduct, or reporting, or dissemination plans of this research.

## Results

Total sample taken from the study location FMRI, Gurugram was 230. Out of this, three participants refused to take part in the study and 71(31.28%) participants were excluded. Reason for exclusion was to have positive troponin value and ST segment changes in ECG. Total 156 participants were included in the study. The mean age of study participants during the time of interview was 44.12 years. Majority of the participants belonged to the age group 31 to 40 years (52, 33.4%). One twenty-four (79.49%) of study participants were male, and 32 (20.51%) were female. Table 1 is showing the sociodemographic characteristics of the participants. Table 2 is showing the distribution of the participants according to various risk factors.

**Table 1:** Sociodemographic characteristics of participants (N=156)

| **Characteristics** | **Male (n=124)**<br>**N (%)** | **Female (n=32)**<br>**N (%)** | **Total N (%)** |
| --- | --- | --- | --- |
| **Age group (years)** |  |  |  |
| 18-20 | 0(0) | 1(3.1) | 1(0.6) |
| 21-30 | 18(14.5) | 7(21.9) | 25(16.0) |
| 31-40 | 46(37.1) | 6(18.8) | 52(33.4) |
| 41-50 | 30(24.2) | 7(21.9) | 37(23.7) |
| 51-60 | 13(10.5) | 8(25.0) | 21(13.5) |
| 61-70 | 6(4.8) | 2(6.2) | 8(5.1) |
| 71-80 | 7(5.7) | 0(0) | 7(4.5) |
| 81-90 | 3(2.4) | 1(3.1) | 4(2.6) |
| >90 | 1(0.8) | 0(0) | 1(0.6) |

**Table 2:** Distribution of participants according to the prevalence of risk factors (self-reported) (N=156)

| **Characteristics** |  | **Male (n=124)**<br>**N (%)** | **Female(n=32)**<br>**N (%)** | **Total N (%)** |
| --- | --- | --- | --- | --- |
| **Smoking** | Present | 37(29.8) | 6(18.8) | 43(27.6) |
|  | Absent | 87(70.2) | 26(81.2) | 140(72.4) |
| **Hypertension** | Present | 28(22.6) | 7(21.9) | 35(22.4) |
|  | Absent | 96(77.4) | 25(78.1) | 121(77.6) |
| **Diabetes** | Present | 13(10.5) | 3(9.4) | 16(10.3) |
|  | Absent | 111(89.5) | 29(90.6) | 140(89.7) |
| **Obesity** | Present | 30(24.2) | 23(71.9) | 53(34.0) |
|  | Absent | 94(75.8) | 9(28.1) | 103(66.0) |
| **Dyslipidemia** | Present | 31(25) | 9(28.1) | 40(25.6) |
|  | Absent | 93(75) | 23(71.9) | 116(74.4) |
| **Past history of CAD** | Present | 16(12.9) | 1(3.1) | 17(10.9) |
|  | Absent | 108(87.1) | 31(96.9) | 139(89.1) |
| **Family history of CAD**<br>**(At age <65 years)** | Present | 6(4.8) | 1(3.1) | 7(4.5) |
|  | Absent | 118(95.2) | 31(96.9) | 149(95.5) |

Fifty participants (32.1%) had diaphoresis, 37 (23.7%) had history of radiation of their chest pain to arms or neck or jaw, 5 (3.2%) had tenderness at the site of their chest pain and 4 (2.6%) had reported worsening in their chest pain with deep inspiration. Fifteen (9.62%) participants reported history of aspirin use within past 7 days but all the participants denied to have history of severe angina ((≥2 episodes in last 24 hours).

Out of 156 participants, 10 (6.41%) reported major adverse cardiac event (MACE) in 30 days of presentation. All the patient reported MACE within 30 days of presentation to hospital with low-risk chest pain were males. All the patients reported MACE within 30 days of presentation to hospital with low-risk chest pain were above 60 years and majority of them were between 71 to 80 years.

Participants who had hypertension reported higher proportion of 30-day Mace (14.29%) in comparison to the participants who did not have hypertension (4.13%).

Participants who had diabetes reported with high rate of 30-day MACE (18.75%) in comparison to the participants who did not have diabetes (5.00%). Participants who were known for coronary artery disease reported with high rate of 30-day MACE (29.41%) in comparison to the participants who were not known for coronary artery disease (3.60%).

Participants who were smokers reported with low rate of 30-day MACE (2.33%) in comparison to the participants who were non-smokers (7.96%), which may be due to most patients with smoking were from young age group and had low prevalence of other risk factors.

Participants who had obesity reported with low rate of 30-day MACE (5.66%) in comparison to the participants who did not have obesity (6.80%) which may be due to presence of higher females was proportionately higher in obese patients. On separate calculation for males and females MACE event rates were higher in smokers than non-smokers.

Participants who had dyslipidaemia reported with high rate of 30-day MACE (15.00%) in comparison to the participants who did not have dyslipidaemia (3.45%). Participants who had family history of CAD at the age <65 years reported with high rate of 30-day MACE (14.29%) in comparison to the participants who did not have family history of CAD at the age <65 years (6.04%).

In this population there was increase in percentage of people who reported 30-day MACE with increase in HEART score. On categorization of HEART score to its low risk (Heart score 0-3) and moderate risk (Heart score 4-6) category as study population did not have participants who can be categorized as per high risk (heart score 7-10); we found that incidence of 30-day MACE was 1.63% in low-risk group (N=123) and 24.24% in moderate risk group (N=33).

With increase in TIMI score in the population there was increase in percentage of people who reported 30-day MACE (Chart 5). On categorization of TIMI score to its low risk (TIMI score 0-1) and moderate risk (TIMI score 2 to 4) category as study population did not have participants who can be categorized as per high risk (TIMI score 5-7), we found that incidence of 30-day MACE was 2.22% in low-risk group (N=135) and 33.33% in moderate risk group (N=21).

With increase in EDACS score in the population there was increase in percentage of people who reported 30-day MACE. On categorization of EDACS score to its low risk (EDACS score 0-15), moderate risk (EDACS score 16-21) and high risk (EDACS score 22 or above); we found that incidence of 30-day MACE was 0.00% in low-risk group (N=126), 14.29% in moderate risk group (N=14) and 50% in high-risk group (N=16).

Main aim of comparing these scores in this study was to find out which score have acceptable incidence of MACE in the low-risk group according to score value in the population. We found that HEART and EDACS score had incidence of MACE less than 2% in their low-risk groups and TIMI score had incidence of MACE >2% in its low-risk group. We also noticed that incidence of 30-day MACE in low-risk group of EDACS score was 0.00% in comparison to low-risk group of HEART score in which incidence of 30-day MACE was 1.63%.

## Discussion

Present study was done to estimate the incidence of 30-day MACE in patients presenting to emergency department with low-risk chest pain. Study population was selected by using non-probabilistic sampling. All the patients presented to emergency department with low-risk chest pain have been included in the study. The diagnosis of low-risk chest pain was done on the basis of absence of ST-segment changes in the ECG on arrival and negatives Zero-hour Troponin-I result. Major adverse cardiac event was identified development of non-ST-elevation myocardial infarction (NSTEMI) or STEMI or emergency revascularization or cardiovascular death or cardiac arrest or cardiogenic shock or high-grade atrio-ventricular block within a 30-day period of presentation to emergency department with low-risk chest pain.

All the 30-day MACE events occurred in males. The probable reason of absence of 30-day MACE events in females is very low number of female participants (32) in the study to detect the 30-day MACE events. Elderly people found to have majority of 30-day MACE events in the present study. Incidence of 30-day MACE was 6.41% in patients presenting to emergency department with low-risk chest pain in this study. Present study finds that hypertension, diabetes, dyslipidaemia, known CAD and family history of CAD were associated with higher incidence of 30-day MACE in patients presenting to emergency department with low-risk chest pain.

In patients presenting to emergency department with low-risk chest pain incidence of 30-day MACE events was 6.41%, which was quite high from acceptable level (<2%). Rainer TH, Leung YK, et al (13) and other various researchers found 30-day MACE events rate ranging from 5 to 15%. Results of our study are similar to these previous studies and this higher than acceptable 30-day MACE incidence validates that it’s not safe to discharge Indian patients with chest pain from emergency department only on the basis of absence of ST-segment changes in the ECG on arrival and negative Zero-hour Troponin-I result.

In present study with increase in HEART score in the population there was increase in percentage of people who reported 30-day MACE. This study also validates that HEART score can be used as a decision-making tool for early discharge tool for patients with low-risk chest pain as there was only 1.63% incidence of 30-day MACE events in participants falling in low-risk category of HEART score (HEART score 0-3).

Backus BE et al (14) showed that incidence of MACE in low-risk category of HEART score was ≤2% with use of HEART score. Results of this study were similar to these validates use of HEART score in our emergency department. In present study with increase in TIMI score in the population there was increase in percentage of people who reported 30-day MACE.

This study also shows that TIMI score is not good enough to be used as a decision-making tool for early discharge tool for patients with low-risk chest pain as there was only 2.22% incidence of 30-day MACE events in participants falling in low-risk category of TIMI score (TIMI score 0-1) which is higher than acceptable level (<2%).

Chase M et al (15) estimated <2% risk in low-risk category (score 0-1) but few studies showed >2% risk with even TIMI score of 1. This study showed result contrast to most studies which may be due to Indian population is different in multiple ways from western population like geographical location, eating habits, exercise habitus etc.

In present study with increase in EDACS score in the population there was increase in percentage of people who reported 30-day MACE.

This study validates that EDACS score can be used as a decision-making tool for early discharge of patients with low-risk chest pain as there was only 0.00% incidence of 30-day MACE events in participants falling in low-risk category of EDACS score (EDACS score <16), which is lower than acceptable level (<2%).

Stopyra JP et al (16) estimated risk of 30-day MACE in low-risk category was <2% with use of EDACS score. This study shows similar results but results to be verified with further studies as this study might not have had enough power to detect MACE events.

This study shows that out of all three scores only EDACS and HEART score can be used as a decision-making tool for early discharge as they had incidence of 30-day MACE within acceptable range (<2%).

EDACS score was best to identify the patients who can be safely discharged from emergency department on presentation with chest pain if they are falling in low-risk group as per EDACS score (30-day MACE incidence 0.00%) but results are to be verified with higher sample size as this study might not have had adequate power to detect MACE events. HEART score was next to EDACS score for the same (30-day MACE incidence 1.63%) and TIMI score was found to be poor in identifying the patients who can be discharged safely from emergency department on presentation with chest pain (30-day MACE incidence 2.22%).

Nieuwets A et al (17) showed that HEART score is better than TIMI score for use as a decision-making tool for early and safe discharge of patients with chest pain from emergency department. This study validates the same finding in Indian patients. There are no studies available for comparison of EDACS score with HEART and TIMI score and this study indicates that EDACS score is better than both HEART and TIMI score for use as a decision-making tool for early and safe discharge of patients with chest pain from emergency department until further comparison high power studies with larger sample size are available.

## Strength and limitations

Study has following strengths. Data collection was done by one investigator. So, there was no inter-observer variation. Response rate was more than 95% (98.7%). Few of the limitations for the study was use of self-reported status for risk factors, this was addressed using verifying the recalled information using probing and checking previous reports or prescriptions. Females were comparatively lower in present study sample (20.5% females compared to 79.5% males). This could limit the generalisability of the result.

## Conclusion

Incidence of 30-day MACE was 6.41% among the patients presenting to emergency department of FMRI Gurugram, Haryana with low-risk chest pain. In low-risk HEART score category 30day MACE was 1.63%, which was lower than acceptable level (<2%). With TIMI score 0 to 1(low risk) associate incidence of 30-day MACE was 2.22% which was higher than acceptable level (>2%). Incidence of 30-day MACE was 0.00% in low-risk category of EDACS score (<16), which was lower than acceptable level (<2%). On comparison EDACS score was best to identify the patients who can be discharged safely from emergency department on presentation with chest pain, HEART score is next for the same but TIMI score cannot be used to do so as with TIMI score 0 to 1(low risk) associate incidence of 30-day MACE was 2.22% which was higher than acceptable level (>2%).

EDACS and HEART score can be used in the Emergency department to identify the low-risk chest pain patients. This could help in early identification and save time and other resources.

## Data Availability

Data will be made available by the corresponding author on reasonable request.

## Acknowledgements

We acknowledge the patients who participated in the study

